# Why Westerners Are Dissatisfied: A Cross-Sectional Study Identifying State-Level Factors Associated with Variation in Private Health Insurance Satisfaction

**DOI:** 10.1101/19004762

**Authors:** Megan McLeod, Jeffrey A. Berinstein, Calen A. Steiner, Kelly Cushing, Shirley A. Cohen Mekelburg, Peter D.R. Higgins

**Author notes:** Corresponding Author: Megan McLeod, BA, 1500 E. Medical Center Drive, Ann Arbor, MI 48109. These authors contributed equally to this work (Megan McLeod, Jeffrey A. Berinstein). Author Contributions: Study concept and design: All authors, Acquisition: Higgins, Berinstein, McLeod, Analysis or interpretation of data: All authors, Drafting of the manuscript: Higgins, Berinstein, McLeod, Figures: Higgins, Berinstein, McLeod, Critical revision of the manuscript: All authors, Final approval: All authors. Disclosures: All authors report no disclosures relevant to this manuscript.

## Abstract

**Importance:** Large regional variations in consumer satisfaction with private health insurance plans have been observed, but the factors driving this variation are unknown.

**Objective:** To identify explanatory state-level and insurance family-level predictors of satsifaction with private health insurance.

**Design:** Cross-sectional study examining regional and state variations in consumer health insurance plan satisfaction using National Committee for Quality Assurance data from 2015 to 2018, state-level health data and parent insurance family.

**Setting:** US Population

**Participants:** Privately insured individuals.

**Exposure:** One of 2176 private health insurance plans.

**Main Outcome:** Consumer satisfaction with the health insurance plan on a 0-5 scale.

**RESULTS:** Consumer satisfaction with health insurance was consistently lowest in the West (*p<*0.0001). Lower private health insurance plan satisfaction was associated with the percentage of the population without a place of usual medical care, the percentage of the state population that is Hispanic, and the percentage of the population reporting any mental illness. Factors associated with increasing insurance satisfaction included higher healthcare spending per capita, a higher number of for-profit beds per capita, and an increased cancer death rate. Increased consumer satisfaction was associated with the Kaiser and Anthem insurance plan families.

**Conclusions and Relevance:** State and insurer family factors are predictive of private health insurance plan satisfaction. Potentially modifiable factors include access to primary care, healthcare spending per capita, and numbers of for-profit hospital beds. This information will help consumers hold insurance providers accountable to provide higher quality and more desirable coverage and provide actionable items to improve health insurance satisfaction.

## Introduction

Insurance plan selection is a complex decision with many important health and financial implications.^1^ Consumers’ ability to make informed decisions about health insurance plans is limited.^2–8^ Gaining insight into the factors contributing to consumer health insurance plan satisfaction will help consumers hold states and insurers accountable and identify actionable items to improve health insurance coverage.

The National Committee for Quality Assurance (NCQA) is an independent nonprofit agency that collects information about non-subsidized health insurance plans and provides both certifications and ratings of consumer satisfaction, prevention, and treatment.^9^ A preliminary analysis of NCQA health insurance satisfaction among private insurance enrollees revealed that there is significant regional and state-level variation in consumer satisfaction (Figure 1). Each state has its own insurance regulations and distinct state-level insurance plans, suggesting that there may be modifiable state-level factors driving consumer satisfaction.

**Figure 1:**
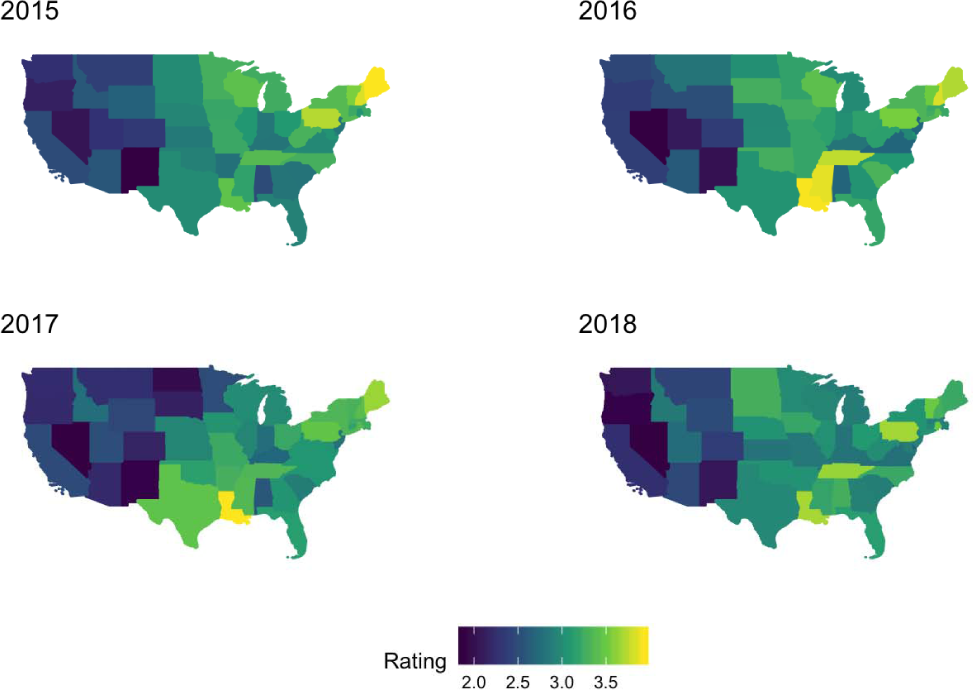
State Variation in Private Insurance Consumer Satisfaction. Maps of the continental United States from the years 2015 to 2018 with mean consumer satisfaction ratings plotted for each state. The mean consumer satisfaction score between Northeast, Midwest, South, and West are significantly different (p<0.0001). Dark blue represents lower consumer satisfaction and yellow represents higher consumer satisfaction. Scores were obtained from the National Committee for Quality Assurance’s (NCQA) Annual Health Insurance Plan Ratings.

This study aimed to identify the regional and state-level factors associated with health insurance satisfaction among private health insurance plan enrollees in the United States. Understanding this variation is the first step toward better meeting the needs of insured individuals.

## Methods

### Study Design

We used a retrospective cross-sectional study design to better understand regional and state-level variations in insurance. Based on our conceptual understanding of insurance plan satisfaction, we extracted data from publicly available repositories for explanatory state-level factors, including demographics, health status, health costs, health providers, and health service utilization. Explanatory independent variables from the year 2016 were used to model the mean state private health insurance plan satisfaction in 2016 NCQA data, as this was the most recent year in which the majority of our candidate explanatory variables were available. We then used this model to make consumer satisfaction predictions for each plan. Institutional review board approval was not obtained for the use of these publicly available aggregate data.

### Data Sources

#### Insurance Satisfaction

We analyzed data from the NCQA’s Annual Health Insurance Plan Ratings from the years 2015 to 2018. The NCQA collects and manages Healthcare Effectiveness Data and Information Set (HEDIS) survey results directly from health plans through the Healthcare Organization Questionnaire, as well as non-survey data from the Interactive Data Submission System. The NCQA’s insurance plan ratings (found at healthinsuranceratings.ncqa.org) are a nationally representative sample of insurance plans that include both accredited and non-accredited health plans.^10^ Ratings are reported on a scale of 0-5 (0 being the lowest and 5 being the highest). Only private (commercial) insurance plans were analyzed. If >10 plans existed in the dataset that shared a common company (i.e. Anthem, Aetna, or Blue Cross), these insurance plans were categorized as an insurance plan family.

### State-Level Predictors

State level predictors were identified using the Kaiser Family Foundation (KFF)’s State Health Facts database, the 2016 United Health Foundation (UHF)’s Annual Report, and the Medical Expenditure Panel Survey (MEPS).^11^ State Health Facts is a project of the KFF that provides health data for all 50 states. Data were extracted directly from the KFF website using C#. Several other sources provided state level factors. We used the 2016 UHF Annual Report to obtain state healthcare quality rankings. The UHF ranks each state across 34 measures of behaviors, community and environment, policy, clinical care, and outcomes.^12^ The percentage of individuals in each state with a high deductible health plan was obtained from MEPS.^13^ Supplemental table 1 provides database details, including a list of variables, variable descriptions, the unit of measure, the year, data source, and the web URL. All of the raw data tables can be found in an Open Science Foundation repository at: https://osf.io/xcbe4/?view_only=06dc35b2496d44f28a2dea4e39bc9dbf.

#### Outcomes and Co-Variables

Our primary outcome was private insurance consumer satisfaction. Insurance satisfaction was treated as a continuous variable and is presented as both a state and regional mean (regions were defined according to the US census definitions as Northeast, Midwest, South, and Southwest). KFF State Health Facts tables were used to identify candidate variables that could serve as mediators of consumer health satisfaction. A full list of exploratory co-variables investigated as well as a web URL for each source data table can be found in Supplemental Table 1.

### Statistical Analysis

Regional variation in the mean consumer satisfaction score was evaluated by analysis of variance (ANOVA). We then developed models using state and the health insurance family as categorical variables to predict the outcome of the mean consumer satisfaction score. Both of these variables contributed significantly to the linear regression model predicton.

To understand the state-level factors driving the association between state and health insurance consumer satisfaction, we then evaluated candidate state-level predictors in a single predictor linear regression model. The promising state-level predictors were then advanced to a multivariable linear regression model to assess for associations between the outcome of private insurance consumer satisfaction and state-level factors, while adjusting for insurance plan family.

Multi-collinearity between predictor co-variables was evaluated visually with a correlogram and by calculation of Pearson correlation coefficients. Collinear independent variables (for example, total hospital beds per capita vs for-profit beds per capita) were selectively excluded from the final regression model, with the better single predictor included in the model. A multivariable linear regression model was then created by forward stepwise inclusion of variables informed by our single predictor models and based our conceptual understanding of insurance plan satisfaction. The variance inflation factor (VIF) was used to examine for multi-collinearity among predictors in our final model. All predictors in the final model had a VIF < 2. Predictors were considered significant if p<0.05 (two-tailed) and the 95% confidence interval (CI) did not include the null hypothesis. The adjusted R-squared was used to estimate the proportion of the variance in consumer satisfaction explained by the state-level factors, while accounting for the number of explanatory variables included in our final model. Subsequently, subgroup analyses of each private insurance plan were performed using multivariable linear regression to examine each private insurance plan separately. All analyses were performed using R version 3.5.0 (R Foundation for Statistical Computing, Vienna, Austria). The corresponding data and analysis code can be accessed in an Open Science Foundation repository at https://osf.io/xcbe4/?view_only=06dc35b2496d44f28a2dea4e39bc9dbf.

### Insurance Plan Performance

We used the explanatory variables from our final multivariable model to make predictions of the expected private consumer insurance satisfaction score. We then subtracted the consumer satisfaction scores predicted by our model from the actual individual consumer satisfaction scores (to obtain model residuals) to identify plans that were over- and under-performing relative to the multivariate model predictions.

## Results

### Private Insurance Consumer Satisfaction

The NCQA’s Annual Health Insurance Plan Ratings for the years 2015 to 2018 included 2176 private (commercial) health insurance plans in total. Satisfaction scores varied significantly by US region for the years 2015 to 2018 (P<0.0001) (**Table 1**). Over these four years, the average private insurance satisfaction score was consistently highest in the Northeast followed by the Midwest (aside from 2017), the South, and lowest in the West. This regioinal difference can be visualized in **Figure 1**. The largest number of state-level predictors were available for the year 2016, so we focused on 2016 data in modeling the outcome of health insurance satisfaction with explanatory state-level and insurance family-level variables.

**Table 1.** Regional Variation in Private Insurance Consumer Satisfaction Score

| Year |  | Northeast<br>(N=466) | Midwest<br>(N=717) | South (N=508) | West (N=485) | Total (N=2176) | p value |
| --- | --- | --- | --- | --- | --- | --- | --- |
| 2015 |  |  |  |  |  |  | < 0.0001 |
|  | Mean Consumer Satisfaction (SD) | 3.3879 (0.3309) | 3.1135 (0.2123) | 3.000 (0.1710) | 2.3800 (0.3283) | 2.9796 (0.4341) |  |
|  | Range | 2.8333 - 3.9286 | 2.5000 - 3.4250 | 2.7857 - 3.4375 | 1.8750 - 3.6000 | 1.8750 - 3.9286 |  |
| 2016 |  |  |  |  |  |  | < 0.0001 |
|  | Mean Consumer Satisfaction (SD) | 3.3291 (0.3030) | 3.1657 (0.2519) | 3.0833 (0.2776) | 2.4057 (0.3843) | 3.0119 (0.4515) |  |
|  | Range | 2.6786 - 3.8000 | 2.6667 - 3.8333 | 2.6765 - 3.8889 | 1.8636 - 3.9000 | 1.8636 - 3.9000 |  |
| 2017 |  |  |  |  |  |  | < 0.0001 |
|  | Mean Consumer Satisfaction (SD) | 3.3448 (0.2165) | 2.9853 (0.3156) | 3.2017 (0.2645) | 2.3664 (0.3929) | 2.9770 (0.4666) |  |
|  | Range | 2.8571 - 3.7143 | 2.0000 - 3.4167 | 2.8125 - 4.0000 | 1.9000 - 4.0000 | 1.9000 - 4.0000 |  |
| 2018 |  |  |  |  |  |  | < 0.0001 |
|  | Mean Consumer Satisfaction (SD) | 3.4144 (0.3121) | 3.2380 (0.2237) | 3.1890 (0.2485) | 2.5256 (0.4751) | 3.1036 (0.4529) |  |
|  | Range | 3.0938 - 3.9250 | 3.0000 - 3.8889 | 2.8125 - 3.9375 | 2.0000 - 4.2000 | 2.0000- 4.2000 |  |

### Factors Associated with Consumer Satisfaction

States as a single predictor accounted for 28.6% of the variance in private insurance consumer satisfaction (Supplemental table 2). Insurance plan family as a single predictor accounted for only 4.63% of the variance in private insurance consumer satisfaction (Supplemental table 3). When controlling for state, several insurance plan families were associated with higher consumer insurance plan satisfaction (Supplemental Table 4) including Anthem (+ 0.5762, 95% CI: 0.2572 to 0.8952, *p=*0.0004), Blue Cross (+ 0.3009, 95% CI, 0.0562 to 0.5456, p=0.0160), and Kaiser (+0.7672, 95% CI, 0.3196 to 1.2147, *p=*0.0008) (Supplemental Table 4). Humana plans were negatively correlated with satisfaction (−0.3552, 95% CI, −0.6636 to −0.0468, *p=*0.0241). States and plan family together accounted for 34.95% of the variance in in private insurance consumer satisfaction.

Using single predictor linear regression, we evaluated associations between our predictor state-level factors of interest and consumer health insurance plan satisfaction. Several state level predictors were significantly associated with consumer satisfaction, and these were advanced to a multivariate model. A complete list of the state-level factors evaluated by single predictor linear regression can be found in Supplemental table 5.

After removing collinear explanatory variables, we constructed a multivariate model to explain the variation in consumer health insurance plan satisfaction with our predictor co-variables while adjusting for insurance plan family (**Table 2**). This model explained 27.78% of the variance in consumer insurance plan satisfaction. The model demonstrates that private insurance satisfaction score decreases by 0.0232 points per each additional percentage of the population reporting having no place of usual medical care (95% CI, −0.0405 to −0.0058, *p=*0.0090), by 0.0222 points per additional percentage of the population who are Hispanic (95% CI, −0.0321 to −0.0124, *p<*0.0001), and by 0.0010 points per each additional percentage of people reporting any mental illness (95% CI, −0.0014 to −0.0006, *p<*0.0001).

**Table 2:** Multivariable Model for Private Health Insurance Satisfaction including State-level Factors and Insurance Family

| Predictor | Regression Estimate (95% CI) | Standard error | P value |
| --- | --- | --- | --- |
| **State-level Factors** |  |  |  |
| Number of for-profit beds (per 1000 persons) | **0.4869 (0.2493, 0.7245)** | **0.1209** | **0.0001** |
| Cancer death rate (deaths per 100,000 persons) | **0.0076 (0.0015, 0.0138)** | **0.0031** | **0.0151** |
| Insurance Dollars (per 1000) spent per capita* | **0.2801 (0.1298, 0.4303)** | **0.0001** | **0.0003** |
| % reporting any mental illness | **-0.0010 (-0.0014, -0.0006)** | **0.0196** | **<0.0001** |
| % of the population who are Hispanic | **-0.0222 (-0.0321, -0.0124)** | **0.0050** | **<0.0001** |
| % reporting no place of medical care* | **-0.0232 (-0.0405, -0.0058)** | **0.8829** | **0.0090** |
| **Parent Insurance plan** |  |  |  |
| Kaiser | **0.7250 (0.2682, 1.182)** | **0.2325** | **0.0019** |
| Anthem | **0.5501 (0.2307, 0.8695)** | **0.1626** | **0.0008** |
| Other | **0.3072 (0.1184, 0.4961)** | **0.0961** | **0.0015** |
| Blue | 0.2368 (-0.0128, 0.4864) | 0.1271 | 0.0630 |
| Cigna | 0.0779 (-0.1659, 0.3216) | 0.1241 | 0.5306 |
| Connecticut General | 0.0787 (-0.1842, 0.3416) | 0.1338 | 0.5567 |
| Coventry | **-0.0720 (-0.3990, 0.2549)** | **0.1664** | **0.6654** |
| Group Health | 0.1925 (-0.2903, 0.6754) | 0.2458 | 0.4337 |
| Health Alliance | 0.3864 (-0.0960, 0.8689) | 0.2456 | 0.1162 |
| Health Net | **-0.0353 (-0.7910, 0.7204)** | **0.3847** | **0.9269** |
| Humana | **-0.2146 (-0.5253, 0.0962)** | **0.1582** | **0.1755** |
| Medica | **-0.0471 (-0.4912, 0.3970)** | **0.2261** | **0.8351** |
| United | **-0.0436 (-0.2453, 0.1580)** | **0.1027** | **0.6710** |

All data are from 2016 unless otherwise stated. \*=data from 2014. Bold font face indicates that the value is statically significant. Red text indicates a negative regression estimate

In contrast, the model demonstrates that private insurance satisfaction score increases by 0.2801 points for every additional 1000 dollars in private health insurance spending per state capita (95% CI, 0.1298 to 0.4303, *p=*0.0003), by 0.4869 points per every additional for-profit hospital bed added per 1000 persons in a state (95% CI, 0.2493 to 0.7245, *p=*0.0001), and by 0.0076 points per every additional cancer death per 100 000 persons (95% CI, 0.0015 to 0.0138, *p=*0.0151).

In this final model, after controlling for state-level factors, three insurance families had significant effects. The Kaiser insurance family added an average of 0.725 points to consumer satisfaction, Anthem added 0.55 points, and small (less than 10 states covered, listed as **Other** in Table 2) insurance plans added 0.30 points of insurance satisfaction on a 5 point scale. No other insurance families had significant estimates in this model. The final model predicts a mean private consumer satisfaction score of 2.75 when all variables in the final model are equal to 0; the crude consumer satisfaction average for 2016 was 3.46. This suggests that these state characteristics are quite important in determining consumer satisfaction with health insurance even after controlling for insurance plan family.

### Insurance Plan Performance

We compared the actual consumer satisfaction score for each of the individual insurance plans to the consumer satisfaction score predicted by our model. **Figure 2** presents the top over-performers and bottom under-performers. A complete list of all 546 insurance plans sorted by the difference between actual performance and predicted consumer satisfaction is presented in Supplemental Table 6.

**Figure 2:**
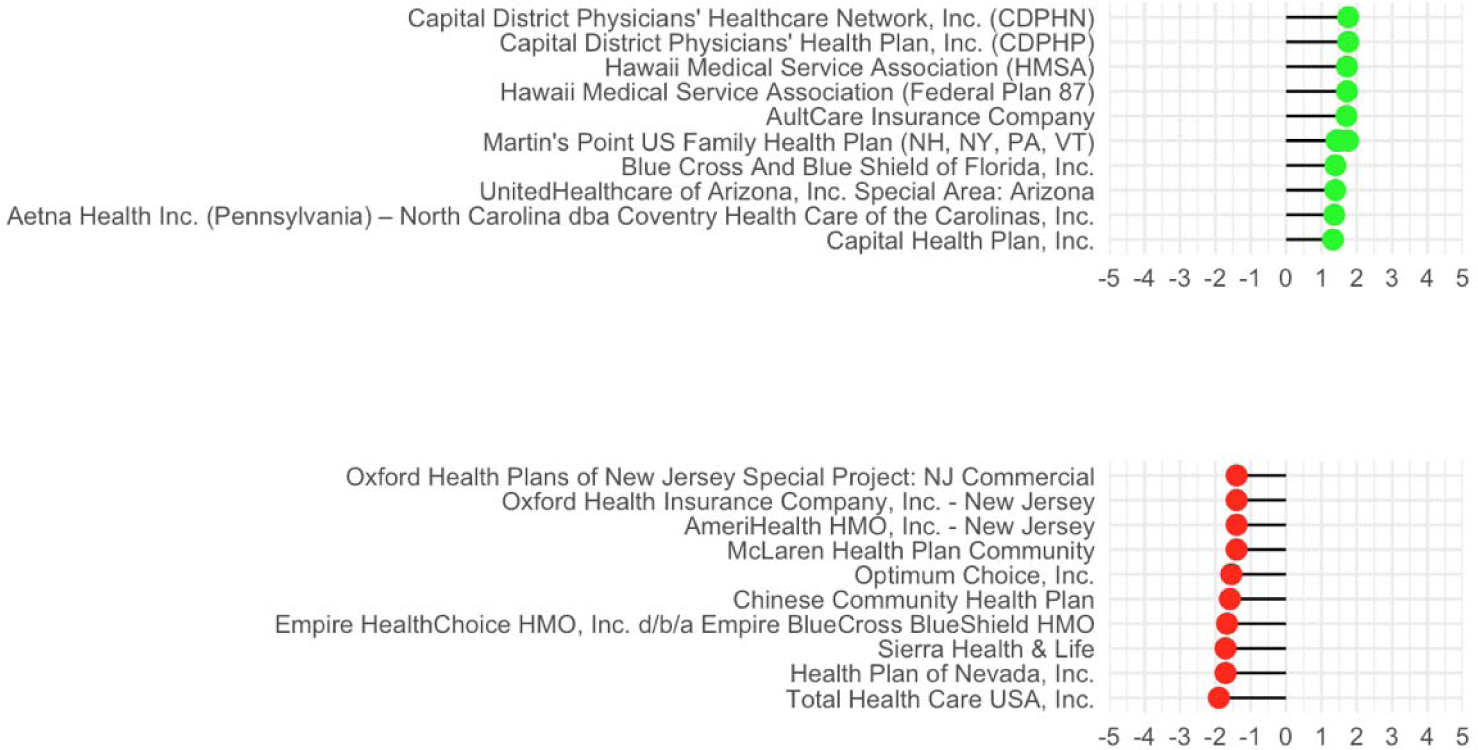
Top Over- and Under-performing Insurance Plans as Predicted by Our Multivariable Model. Residual of the actual consumer satisfaction score obtained by the National Committee for Quality Assurance’s (NCQA) Healthcare Effectiveness Data and Information Set (HEDIS) survey compared to the consumer satisfaction score predicted by our multivariable model which accounts of state as well as various state-level factors.

## Discussion

Large and consistent regional and state-level variations in private health insurance satisfaction in the United States were seen from 2015 to 2018 without an intuitive explanation for low health insurance satisfaction in the western US region. We built a model containing state-level explanatory factors that correlate with this variation with good predictive power. After controlling for insurance plan family, we found that consumer health insurance satisfaction was lower in states that have a higher percentage of individuals without a usual place of medical care, states with a higher percentage of Hispanic individuals, and in states with a higher percentage of individuals reporting mental illness. Conversely, consumer insurance satisfaction was higher in states with higher numbers of for-profit hospital beds, a higher rate of cancer deaths, and higher private insurance spending per capita.

Our finding that states with more people lacking a usual place of medical care have lower insurance satisfaction scores may relate to the notion that those without a primary medical doctor (PMD) rely more heavily on expensive, time-intensive emergency and acute care services for their primary care needs.^14^ Reduced consumer satisfaction among the privately insured population may be mediated by longer wait-times for emergency and acute care services.^15,16^ This finding not only has implications for insurance providers, but also for state and federal government, as previous literature has shown that strong primary care infrastructure results in more equitable health outcomes to society by shifting costs away from more expensive specialty care to less costly primary care through increased resource availability for the most disadvantaged.^17–19^ This also suggests that there are negative externalities for the privately insured when there is a high proportion of individuals without a PMD.

Our finding that states with a higher percentage of Hispanic individuals have lower insurance satisfaction could be in part due to direct effects, as Hispanic individuals experience lower health care satisfaction and lower quality healthcare.^20^ This disparity in care can be partially explained by language barriers, cultural differences, and biases in healthcare access that contribute to general healthcare mistrust among Hispanics.^21–26^ This finding could also be due to indirect effects on the health system, as Hispanics are also more likely to experience barriers to care such as lack of insurance or not having a primary medical doctor, suggesting that this may drive down satisfaction among the privately insured through indirect effects on access to urgent and emergent services as mentioned previously.^21^ As the proportion of Hispanic individuals in the US continues to grow,^30^ particularly in the western US, this could become a larger issue if not adequately addressed.

States with a larger percentage of individuals with mental illness was also associated with lower levels of satisfaction. It has been reported that individuals with severe mental illness have decreased access to routine healthcare and specialty services, and experience a reduced life expectancy of 8-25 years as a result.^31–34^ Disparities in health quality and outcomes are even more striking in ethnic and racial minorities who have a mental disorder.^35–37^ Similar to the logic presented for Hispanic minorities, this finding could be due to direct effects on this vulnerable population, or via indirect effects on the entire health care system which impact all privately insured customers. States and insurers frequently do not adequately meet the needs of people with mental health diagnoses^39,40^ and as a result, mental health care has been identified as an area of critical focus in two recent US Institute of Medicine Reports.^41,42^ This raises the possibility that programs to improve insurance coverage for individuals with mental health disorders could lead to meaningful improvements in access to mental health services and thereby improve consumer satisfaction for not only the mentally ill, but also for all of the privately insured population.

Increased numbers of for-profit beds and per-capita health insurance spending were associated with higher private health insurance consumer satisfaction. Taken together with the association with cancer deaths, these findings suggest that higher health insurance satisfaction may be driven by increased insurance spending. Prior studies have demonstrated that insurance satisfaction is higher among patients with a higher total expenditures and greater prescription drug expenditures.^43^ This is likely to be of particular importance to individuals with chronic conditions as they are likely to receive more healthcare services relative to their financial contribution. In other words, especially high healthcare utilizers could be more satisfied with their health insurance plans because they receive a better return on their investment. It is important to recognize that patient satisfaction does not necessarily correlate with better health outcomes. In addition, the association between increased mortality and consumer satisfaction may reflect a higher mortality in states with more referral centers that are able to provide end stage patients with more aggressive care and even more palliative services at the end of life.

The correlation between consumer health insurance satisfaction and increased healthcare expenditures must be interpreted cautiously. The relationship between treatment intensity and consumer satisfaction poses multiple problems. Although there are multiple studies demonstrating that increased healthcare spending generally leads to better outcomes,^44–48^ untargeted expenditures may have limited marginal health care benefits.^49–51^ While satisfied patients are more likely to be adherent to physician recommendations,^52,53^ patients may request services that are of little or no medical benefit.^54,55^ Physicians may be more inclined to provide these services due to the frequent link between patient satisfaction and physician compensation,^56^ which could have the unintended consequence of iatrogenic harm.^44,45^

Using our findings to identify health insurance plans that are overperforming and underperforming in their state has several important implications for the health insurance market. Consumers can use this information to make more informed decisions about choosing an insurance plan in their area and at the same time hold insurance providers accountable to provide higher quality and more desirable coverage. Insurance companies in turn can use this feedback to continually improve their coverage to meet the needs of their enrollees.

The main limitation of this study is the cross-sectional design, which can only identify associations and cannot deduce causal relationships between these state-level factors and satisfaction among the privately insured. Another limitation is the use of data for private insurance plans, which could have biased our findings so that the number of for-profit hospital beds per capita had a larger effect than total hospital beds per capita. It seems reasonable that the total hospital beds per capita could be a more powerful predictor in a datset that included both public and private health insurance plans.

Additional study limitations include the state-level granularity of the NCQA’s survey data. It is possible that statewide quality measurements with the HEDIS tool may be a crude measure, particularly in large states, as one previous study demonstrated significant geographic variation in plan quality within California.^61^ The main strength of our investigation is the large sample size and large pool of explanatory variables investigated. To our knowledge, the NCQA’s database is the largest health insurance plan satisfaction assessment in the United States, and we believe that this is the best tool currently available for understanding health insurance satisfaction.

These findings also have potential healthcare policy implications. The individual health insurance mandate was repealed as of January 1, 2019, and the Congressional Budget Office (CBO) and the Joint Committee on Taxation (JCT) concluded that the loss of this incentive to enroll could decrease the number of insured persons by 4 million within one year.^57^ Non-subsidized plans are expected to see estimated increases in premiums as high as 16%,^58,59^ highlighting the need for improved understanding of what drives consumer satisfaction with health insurance plans, which in turn contributes to higher enrollment and reduced premiums.^1,60^

The information gained by our study presents several directions for future investigation and potential interventions by insurers and state and federal legislatures. This include (1) increasing the number of people with a primary medical doctor; (2) improving access and care for Hispanic individuals; (3) improving access and care for individuals reporting a mental illness; and (4) increasing the number of for-profit and/or total hospital beds available; All of these interventions could improve consumer satisfaction with health insurance for privately insured patients, and would likely have positive implications for health care quality for uninsured individuals as well.^62^

## Data Availability

All data used in this study are publicly available. All raw and processed data files, as well as code used for study analyses can be found at the link provided.

https://osf.io/xcbe4/?view_only=06dc35b2496d44f28a2dea4e39bc9dbf

## Abbreviations

(ACA): Affordable Care Act
(ANOVA): Analysis of Variance
(HEDIS): Healthcare Effectiveness Data and Information Set
(KFF): Kaiser Family Foundation
(MEPS): Medical Expenditure Panel Survey
(NCQA): National Committee for Quality Assurance’s
(PMD): Primary Medical Doctor
(UHF): United Health Foundation
(VIF): Variance inflation factor

## Notes

Grant Support: MM is supported by TL1TR002242. JAB is supported by T32 DK062708, PDRH is supported by R01 DK109302, R01 DK118154, and T32 DK062708.

### Competing Interest Statement

The authors have declared no competing interest.

### Funding Statement

Grant Support: MM is supported by TL1TR002242. JAB is supported byT32 DK062708, PDRH is supported by R01 DK109302, R01 DK118154, and T32 DK062708.

### Author Declarations

All relevant ethical guidelines have been followed and any necessary IRB and/or ethics committee approvals have been obtained.

Any clinical trials involved have been registered with an ICMJE-approved registry such as ClinicalTrials.gov and the trial ID is included in the manuscript.

